# Sex-Related Differences in Endovascular Treatment Outcomes for Acute Large Infarcts: The ANGEL-ASPECT Subgroup Analysis

**DOI:** 10.1101/2024.11.11.24317138

**Authors:** Dapeng Sun, Xin Guo, Liwen Jiao, Thanh N Nguyen, Mohamad Abdalkader, Yuesong Pan, Mengxing Wang, Gang Luo, Baixue Jia, Xu Tong, Ning Ma, Feng Gao, Dapeng Mo, Raynald, Xiaochuan Huo, Zhongrong Miao, ANGEL-ASPECT study group

## Abstract

**Background and Purpose:** The outcomes of endovascular therapy (EVT) across sexes for large infarcts remains unclear. This study aimed to evaluate the impact of sex on the outcomes of EVT for patients with large infarct.

**Methods:** In this secondary analysis of the ANGEL-ASPECT trial, we compared baseline characteristics and clinical outcomes between men and women, and each cohort further divided into EVT and medical management (MM) groups. The primary outcome was the 90-day modified Rankin Scale (mRS) score distribution. Safety outcomes included symptomatic intracranial hemorrhage (sICH) within 48 h and mortality within 90 days.

**Results:** There were 177 of 455 patients enrolled in the ANGEL-ASPECT trial who were women. 53.7% (95/177) of women and 48.6% (135/278) of men underwent EVT, respectively. The treatment effect of EVT didn’t vary in both sexes with large infarcts (all P >0.05 for interaction). Compared to MM, EVT improved 90-day functional outcomes for both men (3[2-5] vs. 4[3-5], common odds ratio [cOR]: 1.92, 95% CI: 1.26–2.95, P=0.003) and women (4[3-6] vs. 5[4-6], cOR: 2.50, 95% CI: 1.41–4.44, P=0.002). The sICH rate wasn’t different in both treatment groups across both sexes (Men: 5.2% vs. 2.8%, RR: 2.04, 95%CI: 0.56-7.47, P=0.28; Women:7.4% vs. 2.4%, RR:3.00, 95%CI:0.57-15.68, P=0.19).

**Conclusion:** In patients with large ischemic infarct, the treatment effect of EVT didn’t differ between women and men, with better outcomes with EVT versus MM, without an increased risk of sICH. These findings emphasize the need for equal attention and care for both sexes with large infarcts in clinical practice.

## Introduction

Five randomized controlled trials (RCTs) have demonstrated the safety and efficacy of endovascular treatment (EVT) for acute large vessel occlusion (LVO) with large core.^1–5^ The TESLA (Thrombectomy for Emergent Salvage of Large Anterior Circulation Ischemic Stroke trial) did not show a significant benefit of EVT for patients with large infarct.^6^ However, whether sex could modify the outcomes of EVT for large infarct is unknown. While gender equality is increasingly emphasized globally, disparities in real-world settings—such as employment, healthcare, and education—still disproportionately affect women and girls.^7^ Additionally, women may receive less social support and access to rehabilitation than men, which may affect their outcomes after EVT.^8^ Previous studies have investigated the outcome differences of EVT for large vessel occlusion (LVO) without large infarct between men and women, but showed conflicting results and demonstrating worse or neutral outcomes for women compared to men.^9–19^ ^20,21^

As LVO patients with large infarcts often have worse baseline neurological deficit and lower rate of 90-day functional independence after EVT than those without large infarcts, ^22^ it is crucial to further explore these potential inequalities in the context of EVT for large infarcts across different sexes. Therefore, the present study was the secondary analysis of the ANGEL-ASPECT trial to evaluate the influence of sex on EVT outcomes in Chinese patients with large infarcts.

## Methods

### Study Population and Design

This study is a prespecified secondary analysis of the ANGEL-ASPECT trial (NCT04551664),^23^ a multicenter, prospective, randomized, open-label, blinded-endpoint clinical trial conducted in China from October 2, 2020, to May 18, 2022, which enrolled patients with (1) baseline ASPECTS of 3 to 5 on NCCT within 24 hours of symptom onset, (2) baseline ASPECTS of 0 to 2 on NCCT and infarct core volume of 70 to 100 mL within 24 hours of symptom onset, or (3) baseline ASPECTS greater than 5 by NCCT and infarct core volume of 70 to 100 mL between 6 and 24 hours.^3^ Participants were randomly assigned in a 1:1 ratio to either the EVT group or the medical management (MM) group at 46 comprehensive stroke centers. The details of the protocol and primary results of the trial^3,23^ and imaging analyses^24,25^ were reported previously Informed consent was obtained from all patients or their legally authorized representatives prior to enrollment.

### Measurement

Patients were stratified by sex based on the reported data. The baseline, procedural, and outcome variables for this study, including demographic information, premorbid modified Rankin Scale (mRS) scores, mode of admission, medical history, admission National Institutes of Health Stroke Scale (NIHSS) scores, laboratory and imaging results, procedural details, periprocedural management, time metrics, and mRS scores at 90 days, were prospectively collected. The 90-day mRS score was assessed by telephone interview with recording for quality control. An independent imaging core laboratory, blinded to clinical data, reviewed all imaging, including the baseline ASPECTS, occlusion site, recanalization level, target arterial recanalization level at 36 hours, and intracranial hemorrhage. We used baseline CT perfusion or diffusion-weighted imaging (DWI) by RAPID software (Version 5.0.4, iSchemiaView). At least 3 investigators at each site received training and were certified on ASPECTS evaluation. All investigators responsible for enrollment received training on the imaging protocol and use of the RAPID software. Infarct core was defined as the ischemic brain tissue with a relative cerebral blood flow of less than 30% on automated CT perfusion imaging or an apparent diffusion coefficient less than 620 × 10−6 mm^2^/s on DWI.^26^ We assessed the postprocedural arterial recanalization level by expanded Thrombolysis in Cerebral Infarction scale in the EVT group.^27^ Based on the modified arterial occlusive lesion (mAOL) grade on follow-up CT angiography or MR angiography at 36 hours, the degree of recanalization was assessed.^28^

### Study Outcomes

The primary outcome was the 90-day distribution of mRS scores. Secondary outcomes included functional independence, defined as a 90-day mRS score of 0-2, and independent ambulation, defined as a 90-day mRS score of 0-3, early neurological improvement (ENI), measured as a NIHSS score of 0 or 1 or improvement by 10 or more points at 36 hours, change in infarct volume, defined as baseline imaging (CT perfusion or diffusion-weighted imaging) to NCCT at 7 days or at discharge (whichever was earlier) or to MRI at 36 hours, and 36-hour target artery recanalization (mAOL 2-3). ^28^

Safety outcomes included symptomatic intracranial hemorrhage (SICH) and ICH based on the Heidelberg Bleeding Classification within 48 hours, and mortality (mRS 6) within 90 days, and decompressive hemicraniectomy during hospitalization.

### Statistical Analysis

We categorized patients by reported sex into men and women and compared their baseline clinical and imaging characteristics. Further comparisons of baseline clinical and imaging characteristics were made between patients receiving EVT and those receiving MM within each sex group. Continuous variables were reported as medians with interquartile range (IQR), while categorical variables were presented as numbers and percentages. For statistical comparisons, we used the Mann-Whitney test for continuous and ordinal variables, and the Pearson χ2 test or Fisher’s exact test for categorical variables. Baseline characteristics with a p-value <0.1 were selected as the confounders. We applied the ordinal logistic regression model to calculate the common odds ratio (cOR) with the 95% confidence interval (CI) for the ordinal shift in the distribution of the mRS score toward a better outcome for the primary outcome by adjusting for the confounders. The Cox proportional hazards model was used to calculate hazard ratios (HR) with 95% CI for death within 90 days. Moreover, we used a general linear model to calculate mean differences with 95% CI for change from baseline ischemic core volume, and a generalized linear model to calculate relative risk (RR) with 95% CI for other study outcomes. We also investigated whether there was an interaction effect between sex and treatment type on all study outcomes. A two-sided significance level of α=0.05 was applied for all analyses. All statistical analyses were performed using SAS software (version 9.4).

## Results

### Baseline Characteristics

A total of 455 patients were enrolled in the present analysis, of whom 278 were men and 177 were women (Figure 1). Of the 278 men included in this analysis, 135 (49%) underwent EVT, while 95 (54%) of the 177 women received EVT. Baseline clinical and imaging characteristics of the ANGEL-ASPECTS population, stratified by treatment and sex, are summarized in Table 1. Men were younger than women (median [IQR] age 66 [58–72] vs. 71 [64–75] years, p<0.001). Women had a higher prevalence of atrial fibrillation (31.6% vs. 17.3%, p<0.001) and a lower prevalence of current smoking (5.1% vs. 48.6%, p<0.001). Women also presented with more severe strokes, as indicated by a higher median NIHSS score (17 [14–20] vs. 15 [12–19], p=0.009). The distribution of stroke subtypes differed significantly between the sexes (p<0.001), with atherothrombotic strokes more common in men (33% vs. 13.6%) and cardioembolic strokes more frequent in women (57.6% vs. 38.5%). There were no significant differences between the sexes regarding occlusion site, baseline ASPECTS, infarct volume, or workflow times (onset to door, onset to imaging, and onset to randomization). The proportion of patients achieving successful reperfusion was similar between men and women (Table 1 and Table 2).

**Figure 1.**
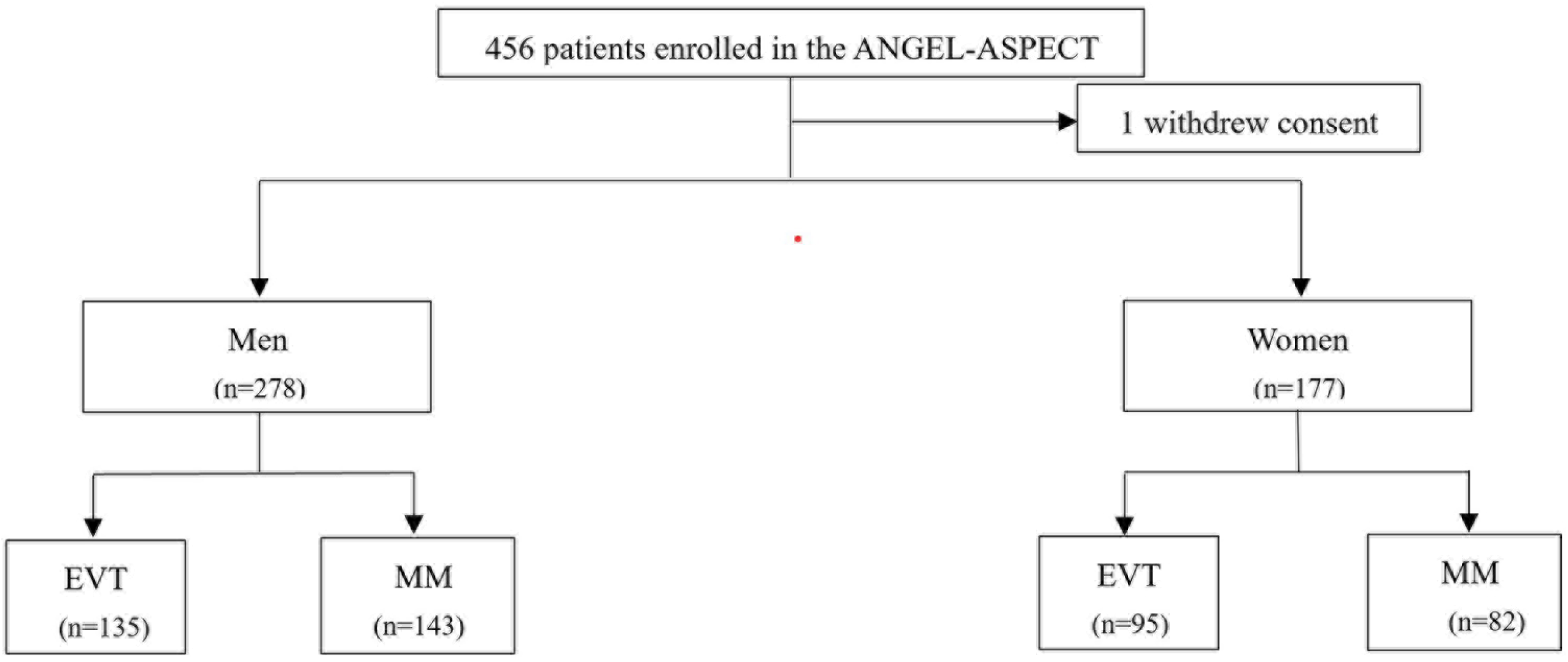
Flow chart. EVT, endovascular therapy; MM, medical management

**Table 1.**
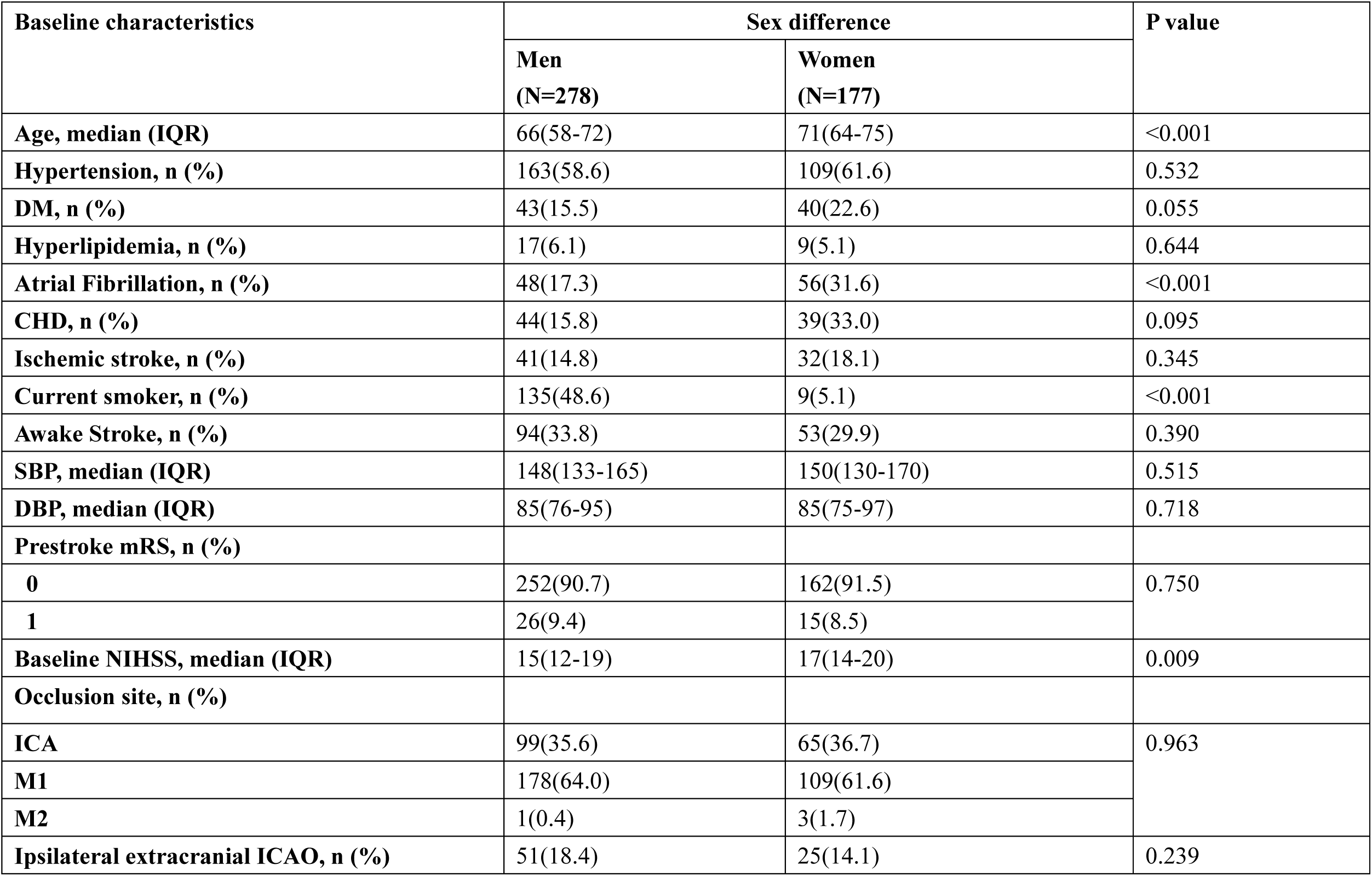
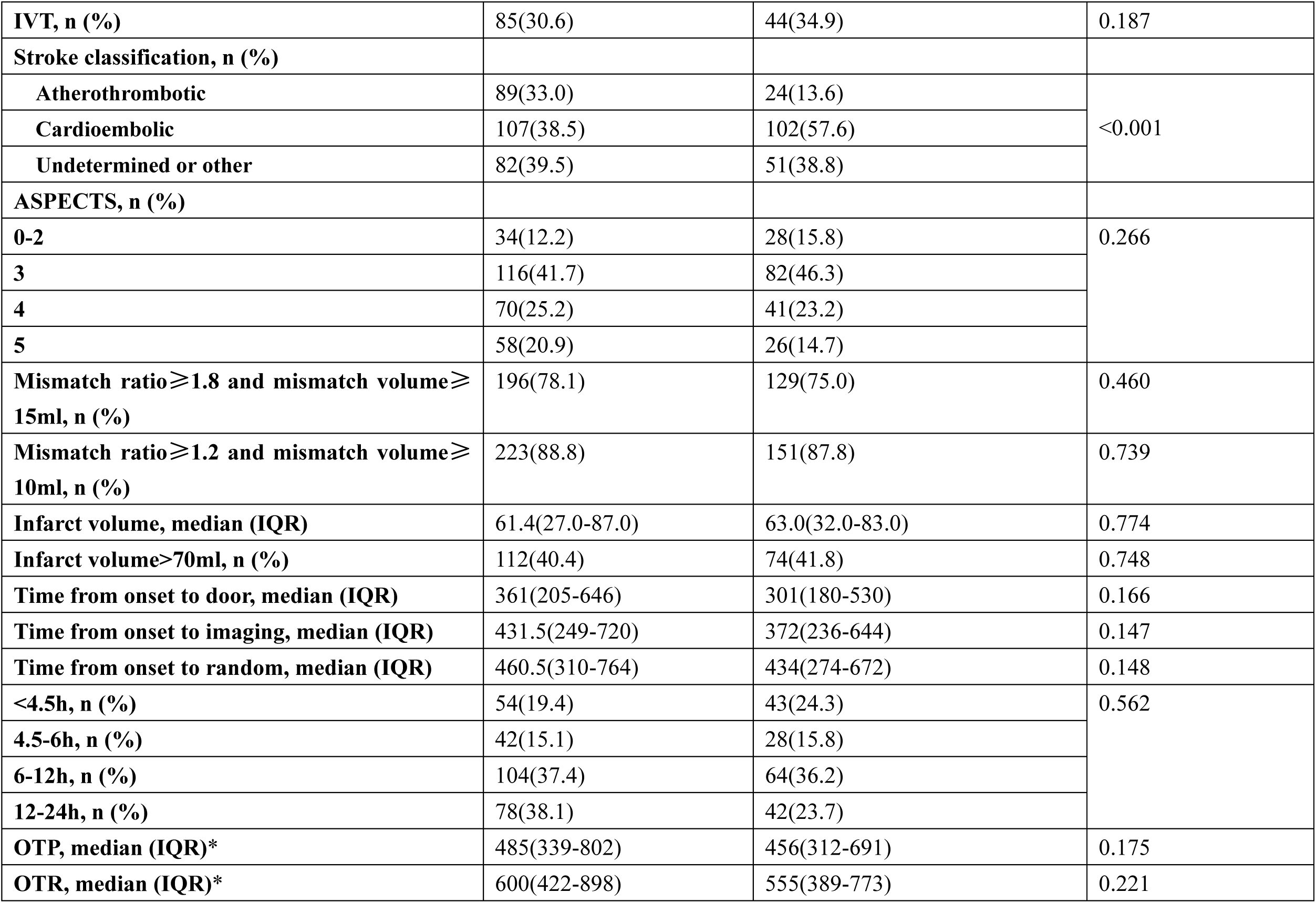
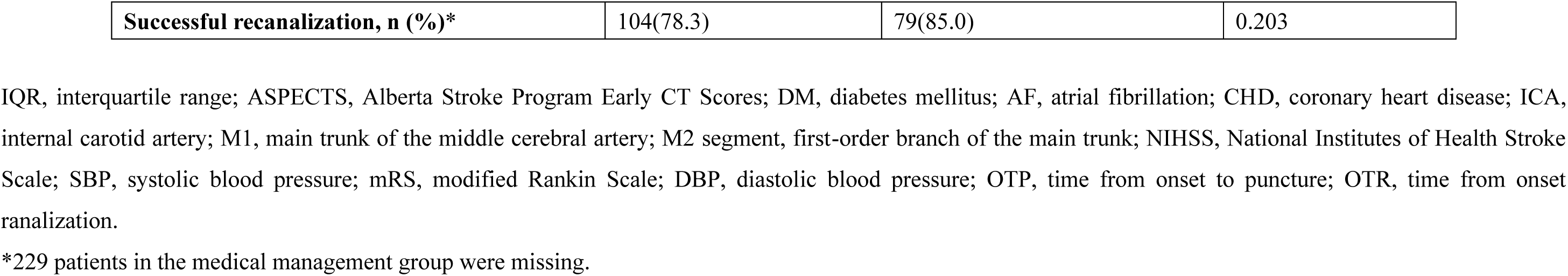
Baseline characteristics between men and women

**Table 2.** Baseline characteristics between EVT and MM in men and women

|  | Sex difference |  |  |  |  |  |
| --- | --- | --- | --- | --- | --- | --- |
|  | Men |  |  | Women |  |  |
| **Baseline characteristics** | EVT<br>(n=135) | MM<br>(n=143) | P value | EVT<br>(n=95) | MM<br>(n=82) | P value |
| **Age, median (IQR)** | 66(58-71) | 67(59-72) | 0.568 | 71(65-75) | 69(61-74) | 0.085 |
| **Hypertension, n (%)** | 76(56.3) | 87(60.8) | 0.442 | 60(63.2) | 49(59.8) | 0.643 |
| **DM, n (%)** | 24(17.8) | 19(13.3) | 0.301 | 19(20.0) | 21(25.6) | 0.374 |
| **Hyperlipidemia, n (%)** | 5(3.7) | 12(8.4) | 0.103 | 8(8.4) | 1(1.2) | 0.039 |
| **Atrial Fibrillation, n (%)** | 21(15.6) | 27(18.9) | 0.463 | 36(37.9) | 20(24.4) | 0.054 |
| **CHD, n (%)** | 18(13.3) | 26(18.2) | 0.268 | 19(20.0) | 20(24.4) | 0.482 |
| **Ischemic stroke, n (%)** | 16(11.9) | 25(17.5) | 0.186 | 21(22.1) | 11(13.4) | 0.134 |
| **Current smoker, n (%)** | 62(45.9) | 73(51.1) | 0.393 | 5(5.3) | 4(4.9) | 0.907 |
| **Awake Stroke, n (%)** | 42(31.1) | 52(36.4) | 0.355 | 27(28.4) | 26(31.7) | 0.634 |
| **SBP, median (IQR)** | 145(130-164) | 151(140-166) | 0.116 | 149(129-170) | 153(130-170) | 0.547 |
| **DBP, median (IQR)** | 85(73-94) | 87(79-96) | 0.159 | 84(73-98) | 87(75-97) | 0.679 |
| **Prestroke mRS, n (%)** |  |  |  |  |  |  |
| **0** | 127(88.8) | 125(92.6) | 0.279 | 78(95.1) | 84(88.4) | 0.110 |
| **1** | 16(11.2) | 10(7.4) |  | 4(4.9) | 11(11.6) |  |
| **Baseline NIHSS, median (IQR)** | 15(12-19) | 15(12-18) | 0.598 | 18(14-21) | 16(13-20) | 0.062 |
| **Occlusion site, n (%)** |  |  |  |  |  | 0.534 |
| **ICA** | 54(37.8) | 45(33.3) | 0.447 | 38(40.0) | 27(32.9) |  |
| **M1** | 88(61.5) | 90(66.7) |  | 55(57.9) | 54(65.9) |  |
| **M2** | 1(0.7) | 0(0) |  | 2(2.1) | 1(1.2) |  |
| **Ipsilateral extracranial ICAO, n (%)** | 27(20.0) | 24(16.8) | 0.489 | 14(14.7) | 11(13.4) | 0.801 |
| **IVT, n (%)** | 38(28.2) | 47(32.9) | 0.393 | 28(29.5) | 16(19.5) | 0.126 |
| **Stroke classification, n (%)** |  |  |  |  |  |  |
| **Atherothrombotic** | 47(34.8) | 42(29.4) | 0.605 | 14(14.7) | 10(12.2) | 0.105 |
| **Cardioembolic** | 49(36.3) | 58(40.6) |  | 60(63.2) | 42(51.2) |  |
| **Undetermined or other** | 39(28.9) | 43(30.1) |  | 21(22.1) | 30(36.6) |  |
| **ASPECTS, n (%)** |  |  | 0.634 |  |  | 0.138 |
| **0-2** | 19(14.1) | 15(10.5) |  | 13(13.7) | 15(18.3) |  |
| **3** | 55(40.7) | 61(42.7) |  | 43(45.3) | 39(47.6) |  |
| **4** | 36(26.7) | 34(23.8) |  | 28(29.5) | 13(15.8) |  |
| **5** | 25(18.5) | 33(23.1) |  | 11(11.6) | 15(18.3) |  |
| **Mismatch ratio $\geq 1.8$ and mismatch volume $\geq 15$ml, n (%)** | 99(78.0) | 97(78.2) | 0.958 | 75(80.7) | 54(68.4) | 0.064 |
| **Mismatch ratio $\geq 1.2$ and mismatch volume $\geq 10$ml, n (%)** | 113(89.0) | 110(88.7) | 0.947 | 85(91.4) | 66(83.5) | 0.117 |
| **Infarct volume, median (IQR)** | 58(24-85) | 65(29-91) | 0.188 | 63(31-90) | 61(32-79) | 0.266 |
| **Infarct volume>70ml, n (%)** | 49(36.3) | 63(44.1) | 0.187 | 41(43.2) | 33(40.2) | 0.695 |
| **Time from onset to door, median (IQR)** | 366(209-646) | 360(181-652) | 0.786 | 320(170-530) | 301(182-659) | 0.718 |
| **Time from onset to imaging, median (IQR)** | 423(247-716) | 438(262-741) | 0.992 | 381(236-643) | 368(231-742) | 0.628 |
| **Time from onset to random, median (IQR)** | 452(306-755) | 469(311-782) | 0.922 | 460(283-669) | 431(261-777) | 0.739 |
| **<4.5h, n (%)** | 25(18.5) | 29(20.3) | 0.855 | 21(22.1) | 22(26.8) | 0.573 |
| **4.5-6h, n (%)** | 20(14.8) | 22(15.4) |  | 16(16.8) | 12(14.6) |  |
| **6-12h, n (%)** | 54(40.0) | 50(35.0) |  | 38(40.0) | 26(31.7) |  |
| **12-24h, n (%)** | 36(26.7) | 42(29.4) |  | 20(21.1) | 22(26.8) |  |
| **OTP, median (IQR)** | 485(339-802) | NA | NA | 456(312-691) | NA | NA |
| **OTR, median (IQR)** | 600(422-898) | NA | NA | 555(389-773) | NA | NA |
| **Successful recanalization, n (%)** | 104(78.2) | NA | NA | 79(85.0) | NA | NA |

IQR, interquartile range; ASPECTS, Alberta Stroke Program Early CT Scores; DM, diabetes mellitus; AF, atrial fibrillation; CHD, coronary heart disease; ICA, internal carotid artery; M1, main trunk of the middle cerebral artery; M2 segment, first-order branch of the main trunk; NIHSS, National Institutes of Health Stroke Scale; SBP, systolic blood pressure; mRS, modified Rankin Scale; DBP, diastolic blood pressure; OTP, time from onset to puncture; OTR, time from onset to recanalization.
\*229 patients in the medical management group were missing.

### Clinical outcomes

Overall, 90-day functional outcomes were improved with EVT in both men (3[2-5] vs. 4[3-5], cOR: 1.92, 95% CI: 1.26–2.95, p=0.004) and women (4[3-6] vs. 5[4-6], cOR: 2.50, 95% CI: 1.41–4.44, p=0.002) (Figure 2), with no significant interaction between sex and treatment (P=0.58) (Table 3). Similar benefits of EVT were observed for functional independence (mRS 0–2) in both men (37.8% vs. 16.1%, RR: 3.84, 95% CI: 2.09–7.05, P<0.001) and women (19.0% vs. 3.7%, RR: 10.22, 95% CI: 2.46–42.40, p=0.001) with no significant interaction (P=0.21). EVT improved independent ambulation (mRS 0–3) in men (57.0% vs. 41.3%, RR: 2.14, 95% CI: 1.26–3.64, P = 0.005) and women (32.6% vs. 19.5%, RR: 3.46, 95% CI: 1.45–8.27, P = 0.005), without significant interaction (P=0.34). Both men (84.0% vs. 40.0%, RR: 9.47, 95% CI: 4.91–18.24, P < 0.001) and women (88.5% vs. 29.7%, RR: 22.08, 95%CI: 7.94–61.39, P < 0.001) demonstrated better target artery recanalization at 36 h in the EVT group than MM group with no interaction (P=0.83). Moreover, ENI was higher in the EVT group than that in the MM group in men (8.2% vs. 2.1%, RR: 4.58, 95%CI:1.22-17.25, P=0.025) but not in women (2.1% vs. 1.3%, RR: 1.13, 95%CI: 0.09-14.09, P=0.923), with no significant interaction (P=0.48) (Table 3).

**Figure 2.**
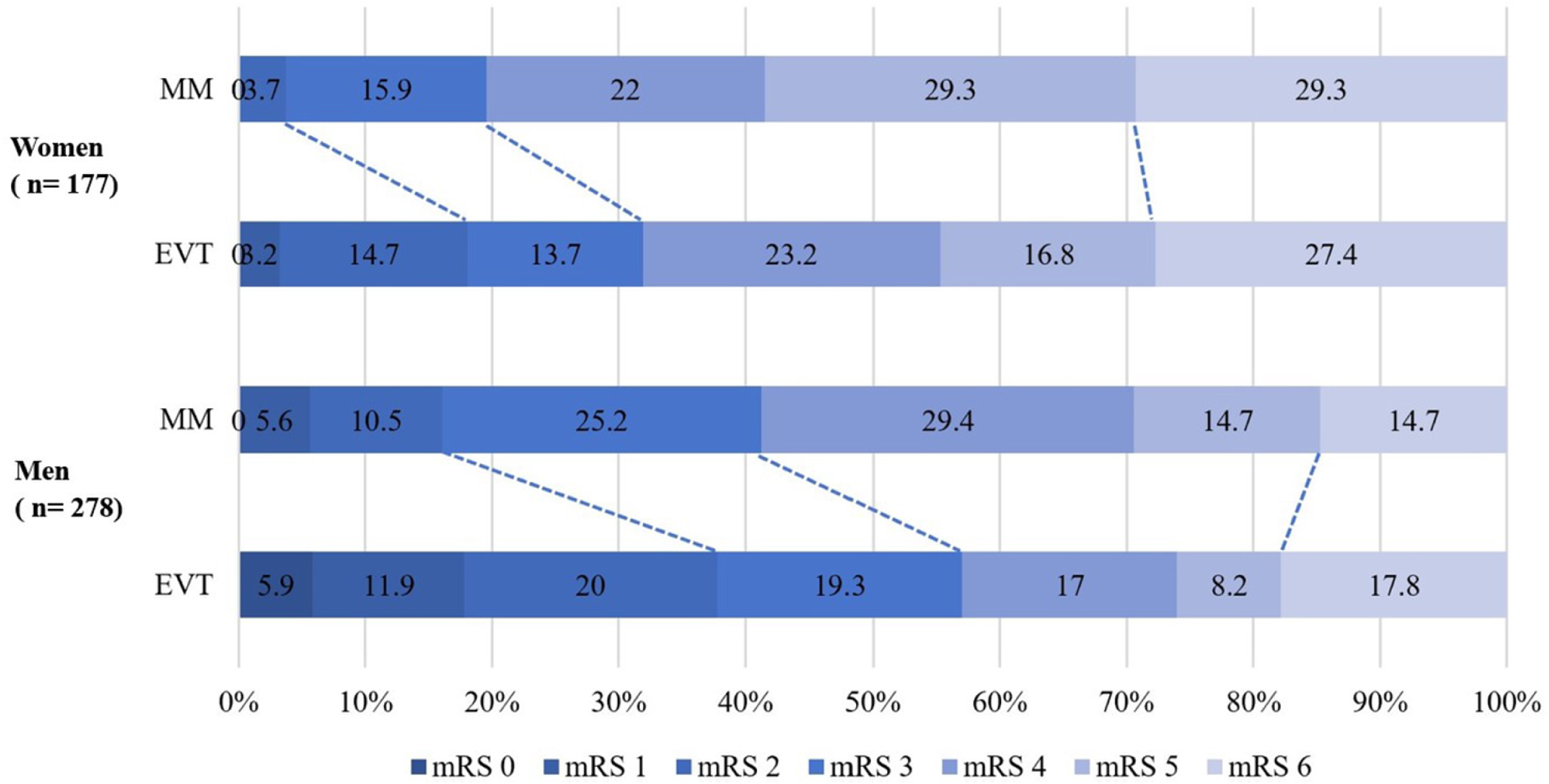
mRS distribution stratified by sex. EVT, endovascular therapy; MM, medical management; mRS, modified Rankin Scale

**Table 3.** Study outcomes

| **Men** | **EVT** | **MM** | **Unadjusted Effect Size\***<br>**95%CI** | **P value** | **Adjusted Effect Size\***<br>**95%CI** | **P value<sup>#</sup>** | **P for**<br>**interaction** |
| --- | --- | --- | --- | --- | --- | --- | --- |
| **mRS, median (IQR)** | 3(2-5) | 4(3-5) | 1.85(1.22-2.82) | 0.004 | 1.92(1.26-2.95) | 0.003 | 0.579 |
| **mRS 0-2, n (%)** | 51(37.8) | 23(16.1) | 3.17(1.80-5.58) | <0.001 | 3.84(2.09-7.05) | <0.001 | 0.205 |
| **mRS 0-3, n (%)** | 77(57.0) | 59(41.3) | 1.89(1.17-3.04) | 0.009 | 2.14(1.26-3.64) | 0.005 | 0.338 |
| **ENI, n (%)** | 11(8.2) | 3(2.1) | 4.11(1.12-15.07) | 0.033 | 4.58(1.22-17.25) | 0.025 | 0.480 |
| **Change volume,**<br>**median (IQR)** | 140.66(63.35-200.43) | 147.28(84.40-227.34) | -14.86(-40.84-11.12) | 0.261 | -14.20(-39.96-11.57) | 0.279 | 0.831 |
| **Target artery**<br>**recanalization at 36**<br>**h, n (%)** | 100(84.0) | 48(40.0) | 7.89(4.28-14.55) | <0.001 | 9.47(4.91-18.24) | <0.001 | 0.156 |
| **SICH, n (%)** | 7(5.2) | 4(2.8) | 1.90(0.54-6.64) | 0.315 | 2.04(0.56-7.47) | 0.282 | 0.771 |
| **Any ICH, n (%)** | 65(48.2) | 26(18.2) | 4.18(2.43-7.19) | <0.001 | 4.81(2.71-8.54) | <0.001 | 0.823 |
| **Death, n (%)** | 24(17.8) | 21(14.7) | 1.11(0.71-1.75) | 0.648 | 1.00(0.61-1.64) | 0.992 | 0.216 |
| **Decompressive**<br>**hemicraniectomy, n**<br>**(%)** | 8(5.9) | 4(2.8) | 2.19(0.64-7.15) | 0.210 | 2.18(0.61-7.73) | 0.229 | 0.824 |
| **Women** | **EVT** | **MM** | **Unadjusted Effect Size\***<br>**95%CI** | **P value** | **Adjusted Effect Size\***<br>**95%CI** | **P value<sup>#</sup>** | **P for**<br>**interaction** |
| **mRS, median (IQR)** | 4(3-6) | 5(4-6) | 1.69(0.99-2.87) | 0.053 | 2.50(1.41-4.44) | 0.002 |  |
| **mRS 0-2, n (%)** | 18(19.0) | 3(3.7) | 6.16(1.74-21.74) | 0.048 | 10.22(2.46-42.40) | 0.001 |  |
| **mRS 0-3, n (%)** | 31(32.6) | 16(19.5) | 2.00(0.99-4.00) | 0.051 | 3.46(1.45-8.27) | 0.005 |  |
| **ENI, n (%)** | 2(2.1) | 1(1.3) | 1.70(0.15-19.09) | 0.668 | 1.13(0.09-14.09) | 0.923 |  |
| **Change volume, median (IQR)** | 131.10(66.04-218.50) | 164.60(101.28-223.10) | -7.63(-37.73-22.48) | 0.618 | -3.60 (-34.32-27.13) | 0.818 |  |
| **Target artery recanalization at 36 h, n (%)** | 69(88.5) | 19(29.7) | 18.16(7.55-43.67) | <0.001 | 22.08(7.94-61.39) | <0.001 |  |
| **SICH, n (%)** | 7(7.4) | 2(2.4) | 3.18(0.64-15.76) | 0.156 | 3.00(0.57-15.68) | 0.193 |  |
| **Any ICH, n (%)** | 48(50.5) | 13(15.9) | 5.42(2.65-11.09) | <0.001 | 5.38(2.50-11.59) | <0.001 |  |
| **Death, n (%)** | 26(27.4) | 24(29.3) | 2.19(0.64-7.45) | 0.210 | 2.18(0.61-7.73) | 0.229 |  |
| **Decompressive hemicraniectomy, n (%)** | 9(9.5) | 4(4.9) | 2.04(0.60-6.89) | 0.251 | 4.17(0.94-18.62) | 0.061 |  |

mRS, modified Rankin Scale; NA, not applicable; CI, confidence interval; ASPECTS, Alberta Stroke Program Early CT Scores; EVT, endovascular therapy; MM, medical management; ENI, early neurological improvement; SICH, symptomatic intracranial hemorrhage; ICH, intracranial hemorrhage.
\* Treatment effects are reported as common odds ratio with the 95% CI for the ordinal shift across the range of mRS toward a better outcome by the method (primary outcome), hazard ratio with the 95% CI for death by a Cox proportional-hazards model, mean difference with the 95% CI for infarct core volume change by the general linear model, and relative risk with the 95% CI for other outcomes by the Cochran–Mantel–Haenszel method.

### Safety outcomes

At 90 days, mortality rates did not significantly differ between the EVT and MM groups in men (EVT: 17.8% vs. MM: 14.7%, hazard risk [HR]: 1.00, 95% CI: 0.61– 1.64, P=0.99) or women (EVT: 27.4% vs. MM: 29.3%, HR: 2.18, 95% CI: 0.61–7.73, P=0.23), with no significant interaction (P = 0.22). Rates of SICH were similar between the EVT and MM groups for men (EVT: 4.2% vs. MM: 2.8%, RR: 2.04, 95% CI: 0.56–7.47, P=0.28) and women (EVT: 7.4% vs. MM: 2.4%, RR: 3.00, 95% CI: 0.57–15.68, P=0.19), with no interaction (P=0.77). The incidence of any ICH was higher in the EVT arm compared to MM in both men (EVT: 48.2% vs. MM: 18.2%, RR: 4.81, 95% CI: 2.71–8.54, P<0.001) and women (EVT: 50.5% vs. MM: 15.9%, RR: 5.38, 95% CI: 2.50–11.59, P<0.001), without interaction (P=0.82). Decompressive hemicraniectomy rates were similar between EVT and MM in men (EVT: 5.9% vs. MM: 2.8%, RR: 2.18, 95% CI: 0.61–7.73, P = 0.229) and women (EVT: 9.5% vs. MM: 4.9%, RR: 4.17, 95% CI: 0.94–18.62, P = 0.061), with no interaction (P= 0.82) (Table 3)

## Discussion

In this secondary analysis of the ANGEL-ASPECT trial, the major findings were that EVT was associated with better clinical outcomes than MM in both men and women without increasing the risk of SICH and death. Additionally, the effect of EVT didn’t vary significantly in both sexes with large infarcts.

Currently, there is no consensus on the benefit of EVT across different sex groups. Although most large-scale studies, such as the HERMES (Highly Effective Reperfusion Evaluated in Multiple Endovascular Stroke trials) patient-level meta-analysis, the SWIFT (Solitaire FR With the Intention for Thrombectomy) and STAR (Solitaire FR Thrombectomy for Acute Revascularization) cohorts, the SOLSTICE (Selection of Late-Window Stroke for Thrombectomy by Imaging Collateral Extent), QuICR (Quality Improvement & Clinical Research) registry, CLEAR (CT for Late Endovascular Reperfusion) study, and ANGEL-ACT (endovascular therapy key technique and emergency work-flow improvement of acute ischemic stroke) registry,^9,16,18,19,29,30^ have shown similar treatment effects of EVT in both men and women, contrasting findings from the MR CLEAN (Multicenter Randomized Clinical Trial of Endovascular Treatment for Acute Ischemic Stroke in the Netherlands) study which suggested better clinical outcomes for men.^10^ This highlights the ongoing uncertainty regarding the differential benefit of EVT by sex. Additionally, there is a lack of evidence on the effectiveness of EVT in patients with large ischemic core. In our study, a secondary analysis of the ANGEL-ASPECT randomized trial, we found that EVT led to better clinical outcomes compared to MM in both men and women at 90 days. However, sex did not influence functional outcomes of EVT at 90-days.

With regards to safety outcomes, mortality did not differ between the EVT and MM groups in either men or women. Although there was a trend toward a higher incidence of sICH and any ICH with EVT in both sexes, the proportion of death was higher in women compared to men. Furthermore, the benefit of EVT over MM for early neurological improvement was more pronounced in men. These findings suggest that women should receive equal attention in stroke units or intensive care units, as studies have shown they are less likely to be treated in these settings—potentially due to misdiagnosis or delayed detection of stroke symptoms.^31,32^ This highlights the importance of national policies that prioritize women’s health, particularly as prior research has indicated that a higher proportion of female patients undergoing EVT live alone and have pre-stroke disabilities, which can delay symptom recognition and emergency response.^14,17,33^

These findings may challenge society to provide greater support to women, as females tend to experience delays in pre-hospital access to care, hospital arrival, access to timely imaging. ^34–36^ Women also experience strokes at an older age compared to males. These sex-related disparities may result in larger infarct strokes in women compared to men. This highlights the need not only for effective stroke management but also for increased focus on timely access to care, post-stroke recovery and rehabilitation, especially given women’s longer life expectancy.^37^ As a result, they may face a higher risk of lacking family or spouse support during recovery. Therefore, further strategies are needed to improve clinical outcomes for women by ensuring more comprehensive support and care during post-stroke management, such as providing community-based support networks, better access to rehabilitation services, and targeted interventions aimed at fostering independence and preventing secondary complications.

While prioritizing women’s health is important, we also highlight opportunity for stroke education and prevention in men based on our findings of a significantly higher rate of smoking use in men. In our cohort, as well as multiple other cohorts evaluating sex differences of stroke in China and Vietnam, there is a consistently higher rate of use of smoking in men compared to women, ^18,19^ which also likely explains the higher rate of intracranial atherosclerotic related large vessel occlusion in men. Therefore, public campaigns and efforts to reduce smoking use in men would be important for primary and secondary stroke prevention.

Our study has several limitations. First, as a subgroup analysis of a RCT, it may be underpowered to fully capture sex-based differences, and the results may not reflect real-world scenarios. However, the use of RCT data allowed us to control for numerous factors that were measured in a consistent, prospective, and rigorous manner. Second, we did not conduct further analysis on early recovery and rehabilitation phase, which may also influence outcomes. Third, we lacked data on sociobehavioral, socioeconomic, and psychological factors, which could influence treatment outcomes between the sexes. Finally, our study population was limited to the Chinese population, which may hinder the generalizability of our results to other ethnic populations.

## Conclusion

In patients with large infarct, the treatment effect of EVT didn’t differ in women and men, with better clinical outcomes with EVT in both groups and without an increased risk of sICH. These findings emphasize the need for equal attention and care for both sexes with large infarcts in clinical practice. A pooled analysis of large core trials stratified by sex strata is warranted.

## Ethics approval

The ANGEL-ASPECT trial was approved by the ethics committee at Beijing Tiantan Hospital (IRB approval number: KY2020-072-02) and all participating centers.

## Author Contributions

DS, XH and ZM contributed to the conception and design of the study; MW and YP contributed to the acquisition and analysis of data; DS and R contributed to drafting the text or preparing the figures. All authors reviewed and approved the final article.

## Data Availability

The data that support the findings of this study are available from the corresponding author upon reasonable request

